# Tennis the sport for life

**DOI:** 10.1101/2025.03.21.25324353

**Authors:** José María Illán Fernández

## Abstract

The aim of the study was to explore the role of tennis in the promotion of health and prevention of disease. The focus was on risk factors and diseases related to a sedentary lifestyle. He registered in PROSPERO (CRD420251016042) a comprehensive literature search was performed across eight databases (PubMed, Scopus, Web of Science, EBSCO, Apunts Medicina de l’Esport, SciELO, MedlinePlus, and Google Scholar) to identify relevant studies published between 2010 and 2025. 17922 articles were found; 18 in relation to the topic we develop in this article, to be studies of either cross sectional or longitudinal design, case– control studies, or experimental studies.

**Objectives:** To evaluate the impact of tennis on life expectancy and its association with the reduction of mortality in different population groups. To investigate the effects of tennis on neuroplasticity and its influence on improving cognitive function, especially in older people. To explore the potential of tennis as a cognitive stimulus for the prevention and rehabilitation of neurological and mental health disorders. To propose practical implications for the incorporation of tennis in public health, active ageing and cognitive rehabilitation programmes.

**Results:** The practice of tennis is associated with cognitive benefits, improvements in physical and bone health. It is also beneficial for people who have suffered from problems with their heart. The methods used to assess these effects vary between the studies reviewed.

From the research it can be concluded that there are very few studies that relate the parameters of physical activities and executive functions, and that there is no heterogeneity between them. In the present study, we aim to explore the association between tennis training experience and executive functions from 4 to 94 years of age, ***tennis the sport for life***.

**Conclusion:** Tennis has a positive impact on overall fitness. Some injuries related to this sport can evolve to chronic in the lower limbs, muscles and tendons, with significant gender differences. Therefore, it is necessary to implement injury prevention strategies.

## INTRODUCTION

Practicing sport contributes to increasing life expectancy, a principle widely supported by doctors, sports science experts, the World Health Organization (WHO), the Academy of Health and Life Expectancy Research, as well as by various biomedical research.

It is worth asking ourselves if all types of exercise generate health benefits? The physical deterioration that occurs over time is associated with a decline in memory and other cognitive functions. Physical activity can attenuate this decline by activating factors that facilitate communication between skeletal muscle and the brain, such as neurotrophins and oxidative stress measurements. It is important to consider that each person’s individual condition influences their life expectancy, in addition to other factors such as pre-existing diseases and physical or biological limitations.

Several studies, which are developed in this article, have shown that tennis produces significant improvements in life expectancy, the so-called “rebound sports”, such as tennis, have the greatest protective effect against cardiovascular diseases.

Tennis is a widely practiced sport worldwide (106 million people according to the 2024 ITF report) that offers physical benefits, but in some cases, it entails suffering injuries due to its claims and its prolonged duration. Tennis-specific injuries are common among professional and amateur players. In addition, scientific statistics link the activity of the sport of tennis with the frequency and severity of injuries.

Tennis is recognized as the sport that contributes the most to the increase in life expectancy, with an average of 9.7 additional years. In addition, it is presented as an effective tool for the prevention and treatment of injuries, which significantly improves the quality of life of those who practice it Paffenbarger R et al. (1).

The practice of tennis causes a notable increase in the entire musculoskeletal system, since this sport is characterized by rapid movements of start and stop, as well as sudden changes of direction. Repeated impacts with the racket also favor the maintenance and increase of muscle mass in the shoulders and arms.

Contemporary tennis technique emphasizes accelerations over direct muscular effort in the arms; This approach is achieved by rotating the hip and shoulders, thus benefiting the core and stabilizer muscles engaged in rotation. Additionally, sprinting, correct posture and sustained effort strengthen the muscles of the legs, back and feet.

The repetitive and continuous movements typical of tennis induce the formation of new bone tissue and facilitate the degradation of aged tissue. The interaction between mechanical stress, hormonal responses, increased blood flow, and increased nutrient absorption is associated with bone strengthening and overall improved skeletal health Babette Pluim (2).

Tennis players have been shown to score outstanding in energy, optimism, and self-esteem, while recording the lowest scores in depression, anger, confusion, anxiety, and tension, compared to other athletes and non-athletes. Since tennis demands alertness and strategic thinking, it fosters new neural connections in the brain, thus promoting continued development in tactics and insight.

Tennis is a sport that can be practiced from the age of four and for a lifetime. People who play tennis periodically per week at a moderate and vigorous intensity cut their risk of death in half, regardless of the cause Paffenbarger R et al. (1).

Regular tennis practice can contribute to the prolongation of the life of practitioners, as long as it is complemented with proper nutrition, regular heart checks, as well as effective sleep and stress management. This sport offers numerous benefits in terms of physical skill and dexterity, as well as increasing both aerobic and anaerobic capacity. However, it is important to consider the impairments associated with tennis, since athletes can suffer injuries that cause a decrease in their performance and, in certain cases, negatively affect their quality of life, even compromising their continuity in this sporting activity, among others García E (3)

### Neuroplasticity and tennis

Brain plasticity was defined in the last century in a pioneering way by William James, which has been updated, among others, by Rodríguez F et al. (4). It is currently understood as the ability of the nervous system to change its structure and functioning throughout its life, as an adaptation to the diversity of the environment, at the same time, strong enough that it is not completely modified. A concept widely used in neuroscience and psychology in a non-peaceful way.

Due to neuroplasticity, neurons are regenerated both anatomically and functionally and form new synaptic connections. On the other hand, what is known as neuronal plasticity represents the brain’s ability to recover from damage and restructure itself.

Spatial cognition facilitates the successful performance of specific cognitive tasks through lateral processing and neuroplasticity. Long-term training in tennis induces efficiency in the neural processing of the visuospatial cognitive cortex of athletes. However, the lateralization features and neural mechanisms of visuospatial cognitive processing in tennis players in non-sports settings are unclear, among other authors Peng, Z et al. (5)

### Tennis as a cognitive stimulus

Cognitive stimulation is another expected outcome. Playing tennis can: improve executive functions (such as working memory and decision-making), reduce the risk of age-related cognitive decline such as Alzheimer’s or dementia, increase brain plasticity, which implies that the brain maintains its ability to reorganize and adapt to new tasks or demands.

Tennis requires quick decision-making, visuospatial coordination, and sustained attention, making it an excellent cognitive stimulus. This is supported by studies such as that of Peng et al. (7) in tennis players (comparable model), which show changes in brain structure associated with sports practice.

## METHOD

The objective of this strategy was to identify, select and review relevant and high-quality studies that evaluated the impact of tennis on life expectancy, neuroplasticity and cognitive stimulation. The search focused on clinical trials, observational studies, systematic reviews and intervention studies in indexed scientific databases.

### Eligibility Criteria

To be included in this research, studies had to meet the following criteria:

### Sources of Information

The following databases will be used for the search: PubMed, Scopus, Web of Science, EBSCO, Apunts Medicina de l’Esport, SciELO, MedlinePlus, and Google Scholar.

### Keywords

The search used MeSH terms by combining them with key terms and Boolean operators. Some of the key terms and operators used were: **Terms MeSH:** tennis, physical, activity, exercise, Neuroplasticity, cognition, life expectancy, cognitive function, aging, sports medicine. **Additional Keywords:** impact of tennis on life, expectancy, neuroplasticity and tennis, cognitive benefits of tennis, longevity and physical activity, exercise and brain function, tennis and brain health, sport and cognitive aging. **Boolean Operators:** AND (para combinar términos relacionados), OR (para incluir sinónimos o términos relacionados), NOT (para excluir términos irrelevantes).

### Inclusion and exclusion criteria

**-Inclusion criteria:** studies published between 2010 and 2025, peer-reviewed articles (scientific journals), studies focused on ages 4 to 94, research on tennis and its impact on physical health, neuroplasticity, and cognitive function, clinical trials, cohort studies, observational studies, systematic reviews, publications in English.
**-Exclusion criteria:** non-tennis-related study, studies without information on neuroplasticity or cognitive function, studies in pediatric populations or with advanced neurodegenerative diseases, articles not peer-reviewed or without methodological rigor, publications in languages not accessible (e.g., non-translatable).

Detailed Search Strategy PubMed:

1. (“Tennis”[MeSH] OR “tennis”[TI]) AND (“Neuroplasticity”[MeSH] OR “brain plasticity”[TI]) AND (“Cognitive Function”[MeSH] OR “cognition”[TI]) AND (“Life Expectancy”[MeSH] OR “longevity”[TI])
2. (“Tennis”[MeSH] OR “tennis”[TI]) AND (“Physical Activity”[MeSH] OR “exercise”[TI]) AND (“Cognitive function”[MeSH] OR “neuroplasticity”[TI]) AND (“elderly”[TI] OR “aged”[MeSH])
3. (“Neuroplasticity”[MeSH] OR “brain plasticity”[TI]) AND (“Tennis”[MeSH] OR “physical activity”[TI]) AND (“Cognition”[MeSH] OR “mental health”[TI])

Scopus:

1. TITLE-ABS-KEY (“Tennis” AND “Neuroplasticity” AND “Cognition”) AND (LIMIT-TO (DOCTYPE, “ar”))
2. TITLE-ABS-KEY (“Life Expectancy” AND “Exercise” AND “Cognitive Function”) AND (LIMIT-TO (DOCTYPE, “ar”))
3. TITLE-ABS-KEY (“Tennis” AND “Brain Health” AND “Longevity”) AND (LIMIT-TO (DOCTYPE, “ar”))

Web of Science:

1. TS= (“Tennis” AND “Neuroplasticity” AND “Cognitive Function”)
2. TS= (“Physical Activity” AND “Tennis” AND “Life Expectancy”)
3. TS= (“Exercise” AND “Brain Function” AND “Neuroplasticity”)

SPORTDiscus:

1. “Tennis” AND “Neuroplasticity” AND “Cognitive Aging”
2. “Tennis” AND “Longevity” AND “Physical Activity”
3. “Cognitive Function” AND “Exercise” AND “Brain Plasticity”

#### Study Selection Strategy

**– Review of Titles and Abstracts:** Initial searches of databases will be carried out and the titles and abstracts of the studies found will be reviewed. Those that meet the inclusion criteria will be selected.
**– Full Review of Articles:** Articles that pass the title and abstract review phase will be read in their entirety to verify that they meet all inclusion criteria.
**– Final Selection:** The selected articles will be organized into categories according to the type of study: clinical trials, observational studies, systematic reviews, etc.
– Data Analysis and Extraction

For each selected study, the following data will be extracted:

- Author information and year of publication.
- Objective of the study and type of design (clinical trial, cohort study, review, etc.).
- Study population (age, sex, characteristics of participants).
- Intervention or exposure (tennis and other forms of exercise).
- Outcome measures (life expectancy, neuroplasticity, cognitive function).
- Main findings and conclusions of the study.

#### Assessment of the quality of Studies

Tools such as the Observational Study Quality Scale (STROBE) were used to assess the methodological quality of the selected studies. Clinical trials were evaluated using the PEDro Scale.

#### How to merge data

- **Selection of relevant studies**: Studies were selected based on inclusion and exclusion criteria and grouped according to their type (quantitative or qualitative).
- **Qualitative synthesis**: For qualitative studies, a narrative synthesis was carried out by grouping and comparing the relevant findings.
- **Calculation of combined effects**: For quantitative studies, data from selected studies were combined using a meta-analysis
- **Assessment of heterogeneity**: we assessed heterogeneity between studies to determine whether it is appropriate to combine results. Using I² statistics and heterogeneity tests.

## Results

Tennis is associated with a longer life expectancy. Robust research Schnohr et al.; Oja et al.; Paffenbarger et al. (6)(8) (1) show that sports such as tennis are associated with a reduced risk of mortality and a significant increase in longevity (up to +9.7 years according to the Copenhagen City Heart Study).

Playing tennis regularly can be an effective cardiovascular prevention and health promotion strategy.

Tennis requires quick decision-making, visuospatial coordination, and sustained attention, making it an excellent cognitive stimulus. This is supported by studies such as that of Peng et al. (5) in table tennis players (comparable model), which show changes in brain structure associated with sports practice.

Tennis could be useful as a cognitive training tool in healthy people or even in active aging. There is a theoretical consensus García, James W (3) (7); informative articles) in which activities such as tennis stimulate the release of factors such as BDNF, which promote brain plasticity and learning.

Tennis may help maintain brain health, although more direct experimental studies are needed. Although they are not part of the indexed articles, many popular texts emphasize that tennis improves mood, reduces stress and strengthens social bonds. Tennis has potential as an integral tool for physical, mental and social well-being.

Figures and Tables: continued on the following pages.

### PRISMA Flow Chart

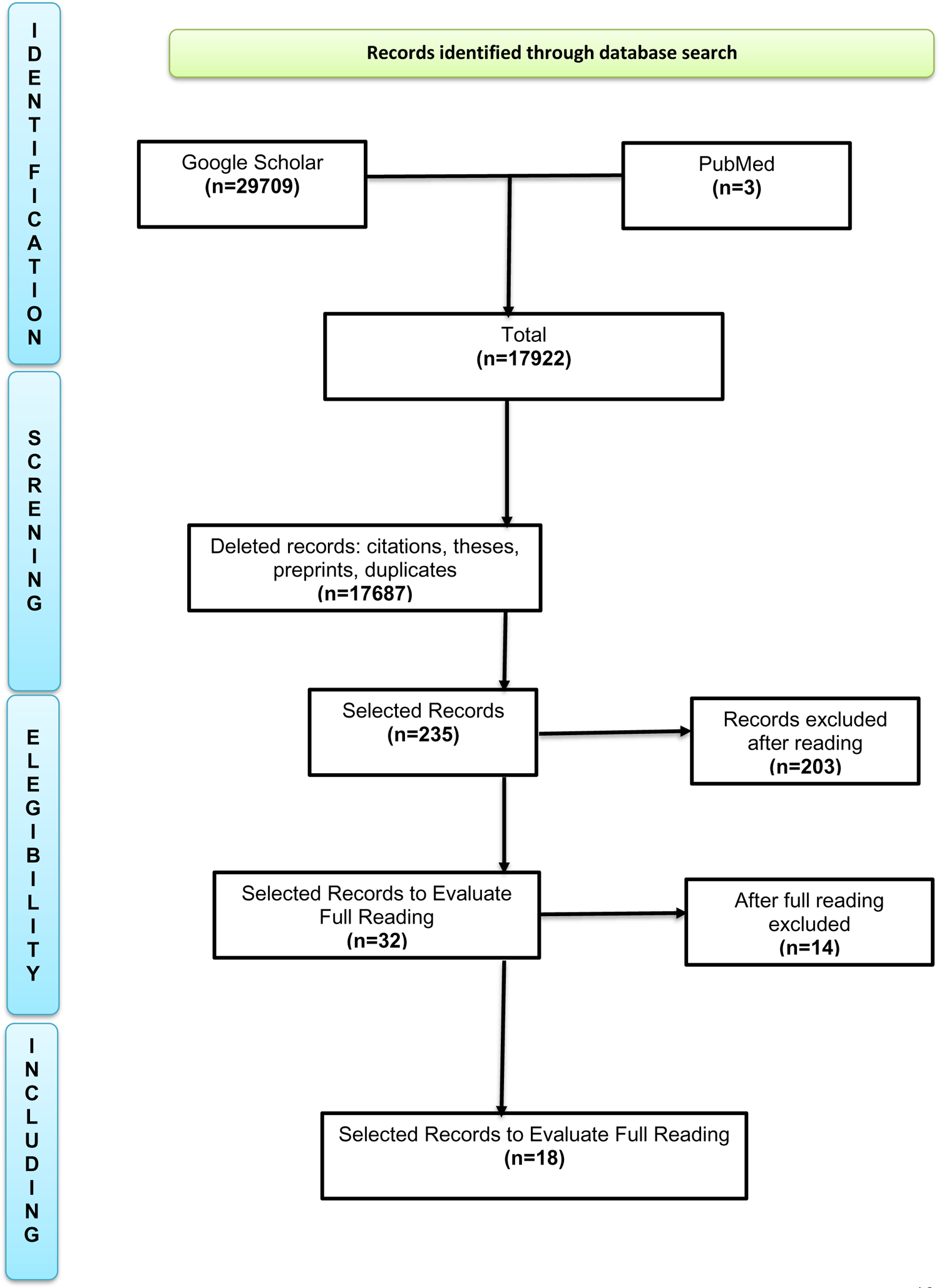

**Table 1.** Assessment of the quality of the evidence (GRADE), La **Tabla 1**, reflects the values of asssessment of the quality of the evidence (GRADE), of this research work. High: High confidence that the estimated effect is close to the real one. Moderate: Moderate confidence; The actual effect may be different. Low: Limited evidence, susceptible to new findings. BDNF: Brain-Derived Neurotrophic Factor, key in synaptic plasticity. Which we can summarize in **table 2**.

| Thematic area | Type of studies | Initial design | Consistency | Precision | Direction of the effect | Overall quality of the evidence |
| --- | --- | --- | --- | --- | --- | --- |
| Life expectancy | Estudios de cohortes longitudinales (Schnohr et al., 2018; Oja et al., 2017; Paffenbarger et al., 1986) | Loud | High (replicated results) | Good (narrow CIs) | Positive | Loud |
| Neuroplasticity and cognition | Narrative review, experimental studies (table tennis), popular science articles | Moderate | Moderate (consistent findings) | Limited (few direct studies) | Positive | Moderate |
| Physiological mechanisms (BDNF, lateralization, decision-making) | Theoretical and experimental studies (Peng et al., 2022; William J., Garcia, Schwartz, Holmes) | Moderate | Moderate | Variable | Positive | Moderate-low |

**Tabla 2.** Possibility of applying the findings of this research.

| Category | Level of evidence | Applicability |
| --- | --- | --- |
| Life expectancy | Loud | Recommended |
| Cognitive stimulation | Moderate | Recommended |
| Neuroplasticity | Moderate/Low | Hypothesis |
| Emotional well-being | Theoretical | Possible |

## DISCUSSION

Tennis, as a physical activity, has proven to be a practice with multiple health benefits, which go beyond simple physical conditioning. Various studies suggest that this sport can positively influence life expectancy, neuroplasticity and cognitive development, which highlights its value in the field of public health and prevention.

Several studies Paffenbarger et; Schnohr et al., 2018; Oja et al., 2017) (1) (6)(8) have documented that regular tennis practice is associated with greater longevity. These studies confirm that people who participate in intense physical activities, such as tennis, have a higher chance of surviving cardiovascular disease and other conditions related to a sedentary lifestyle. The analysis of the relationship between physical activity and longevity is well established in the literature, but tennis, being a sport that combines aerobic and anaerobic exercise, seems to be particularly effective in reducing mortality from general and cardiovascular causes. However, more direct intervention studies that can isolate the specific effects of tennis versus other forms of exercise are needed to support this conclusion Oja, P. et al. (8).

Studies on neuroplasticity reveal that playing tennis, like other sports that require coordination, agility and quick decision-making, can induce changes in brain structure and function. Research such as that of Peng et al. (5) on tennis has shown that this type of sport can promote the lateralization of spatial cognition and improve executive function, particularly in the dominant hemisphere. Although this finding is promising, it is important to note that direct evidence on tennis-specific induced neuroplasticity remains limited and needs further experimental exploration. In addition, most studies focus on related sports, such as table tennis, which raises the need for studies more focused on tennis itself.

The impact of tennis on mental health is another relevant aspect. Physical activity in general is associated with improved mood, stress reduction, and prevention of disorders such as depression and anxiety. Unlike other sports, tennis demands strategy, concentration and adaptability in high-pressure situations, which contributes to mental well-being. Although experimental studies are limited, popular articles García (3) point out that tennis, as a social sport, also facilitates interaction and a feeling of belonging, important factors for emotional health. Despite these theoretical benefits, it is critical to consider individual variability in the emotional effects of playing tennis, as some individuals may experience elevated levels of frustration or stress in their learning and competition.

### Limitations in research

Although the literature on tennis and health is promising, there are also some limitations to the current research. Much of the evidence comes from observational studies that do not allow direct causality to be established, suggesting the need for controlled clinical trials that more rigorously explore the specific effects of tennis in different populations (e.g., older adults or people with neurological conditions). In addition, many of the studies reviewed focus on racquet sports in general, making it difficult to generalize the results exclusively to tennis.

The quality of studies on neuroplasticity also varies, and much of the evidence comes from theoretical and experimental work related to table tennis, underscoring the importance of having studies that directly focus on the relationship between tennis and brain changes.

### Recommendations, future research and clinical application

The results of the studies reviewed open the door to new areas of research. There would be value in conducting randomised controlled trials specifically examining the effects of tennis on cognitive function and neuroplasticity, particularly in older people or those at risk of neurodegenerative diseases. In addition, it would be interesting to investigate how different intensities and types of tennis training (singles vs. doubles, recreational vs. competitive) can influence physical and mental health outcomes.

In terms of clinical application, tennis can be incorporated into cardiovascular disease prevention and rehabilitation programs, as well as cognitive stimulation interventions for the elderly or people with neurological conditions. The combination of physical and mental exercise in tennis is one of its advantages, as it requires both physical and cognitive skills.

In short, tennis presents multidimensional benefits including improving life expectancy, stimulating neuroplasticity, and promoting emotional and social well-being. Si bien la evidencia respalda en gran medida estos efectos, se necesita mayor investigación para consolidar estos hallazgos, especialmente en lo que respecta a la neuroplasticidad y los mecanismos cognitivos inducidos por la práctica del tenis.

## CONCLUSIONES

Currently, the evidence linking tennis practice with an increase in life expectancy comes mainly from observational studies, such as the Copenhagen City Heart Study (6), which showed a significant association between the practice of racket sports and greater longevity. However, no randomised clinical trials (RCTs) have been identified that confirm this causal relationship. RCTs are considered the gold standard for establishing causal relationships, but their implementation in this context is complex due to ethical and logistical factors.

Importantly, although observational studies suggest a strong association between tennis practice and longer life expectancy, the absence of RCTs implies that we cannot say with certainty a direct causal relationship.

Further research, preferably with experimental designs, is required to confirm these findings and better understand the underlying mechanisms.

Life expectancy and tennis: high-quality evidence based on observational criteria, but no randomised trials.

Neuroplasticity and tennis: moderate, promising but still emerging and somewhat indirect evidence.

### Rationale for this research

Physical activity has been widely recognized as a crucial factor for improving health and preventing chronic diseases, including cardiovascular disease, diabetes, and neurodegenerative disorders. However, among the various forms of exercise, tennis has received less attention in the scientific literature compared to other more popular sports such as running or swimming, despite its potential benefits for physical, mental and cognitive health. This article aims to analyse the specific effects of tennis practice on life expectancy, neuroplasticity and cognitive stimulation, and to offer a comprehensive view of the benefits that this sport can bring to the population.

While there is research highlighting the positive effects of physical exercise on health, few studies focus on tennis and its unique impact on long-term health. According to the available evidence, sports such as tennis can not only increase longevity and reduce the risk of mortality from cardiovascular disease, but also have positive effects on brain function. Current research on neuroplasticity and cognitive training related to tennis is limited and therefore there is a gap in knowledge that needs to be addressed.

In the context of aging, where the maintenance of cognitive health is a priority, tennis offers a promising alternative not only for the preservation of physical health, but also for the promotion of mental activity. Studies on neuroplasticity suggest that activities such as tennis can modify brain structure, enhancing the brain’s ability to adapt and learn. This is especially relevant for people who are elderly or at risk of developing neurodegenerative disorders.

On the other hand, the social and emotional component of tennis, being a sport that can be played as a couple or in a group, also provides benefits in terms of stress reduction, improved emotional well-being and promotion of social interaction. These aspects have important implications in improving quality of life and preventing mental illnesses, such as anxiety and depression.

This work aims to fill the gap in the scientific literature on the effects of tennis, offering a more complete view of how this sport can contribute to both the improvement of life expectancy and cognitive stimulation. By integrating neuroplasticity into the analysis, a novel approach is on the brain and cognitive function. In addition, it is argued that tennis could be a useful tool in prevention and rehabilitation programs, especially in older people or people with neurological conditions.

In practical terms, this article could have important implications for healthcare professionals, such as doctors, physiotherapists and psychologists, who are looking to incorporate physical and cognitive activities into their intervention programmes. In addition, public policies on health and active aging could consider tennis as a viable option to promote a healthy and active lifestyle in the population.

This article provides a comprehensive approach that combines the evidence on the benefits of tennis with the concept of neuroplasticity, an area not yet fully explored in relation to this sport. Through a review of the literature, it is highlighted how tennis, beyond being a simple form of exercise, can be a powerful cognitive stimulant, contributing to the physical and mental well-being of individuals.

With the increasing prevalence of neurodegenerative diseases and emotional well-being disorders in ageing societies, tennis could play a key role in prevention, rehabilitation and improved quality of life. In addition, the article provides a practical and applicable analysis that may be useful for the formulation of public health programs and clinical intervention strategies.

In summary, our research contributes to recognizing tennis as a multidimensional tool that promotes both physical and cognitive health, and opens the door for future research that delves into these effects and the best ways to apply this sport in preventive and therapeutic contexts.

## Data Availability

All data produced in this work are contained in the manuscript

## ACKNOWLEDGEMENTS

I want to express my sincere thanks to all the people and institutions that made it possible to carry out this research. First of all, I am deeply grateful to Dr. Fernando Jiménez Conde, director of my doctoral thesis, for his invaluable guidance, support, and patience throughout this project. Their experience and knowledge were fundamental for the development of my research.

## CONFLICT OF INTEREST AND FUNDING

The author declares that he has no conflict of interest and that they have not received funding to carry out the research.

## Notes

### Competing Interest Statement

The authors have declared no competing interest.

### Funding Statement

without financing

